# Computationally-informed insights into anhedonia and treatment by *κ*-opioid receptor antagonism

**DOI:** 10.1101/2024.04.09.24304873

**Authors:** Bilal A. Bari, Andrew D. Krystal, Diego A. Pizzagalli, Samuel J. Gershman

## Abstract

Anhedonia, the loss of pleasure, is prevalent and impairing. Parsing its computational basis promises to explain its transdiagnostic character. We argue that one manifestation of anhedonia— reward insensitivity—may be linked to limited memory capacity. Further, the need to economize on limited capacity engenders a perseverative bias towards frequently chosen actions. Anhedonia may also be linked with deviations from optimal perseveration for a given memory capacity, a pattern that causes *inefficiency* because it results in less reward for the same memory cost. To test these hypotheses, we perform secondary analysis of a randomized controlled trial testing *κ*-opioid receptor (KOR) antagonism for anhedonia, as well as analyses of three other datasets. We find that anhedonia is associated with deficits in efficiency but not memory, whereas KOR antagonism (which likely elevates tonic dopamine) increases memory and efficiency. KOR antagonism therefore has distinct cognitive effects, only one related to anhedonia.

## Introduction

Anhedonia, the loss of pleasure or lack of reactivity to pleasurable stimuli, is observed in many psychiatric illnesses, including major depressive disorder, bipolar disorder, schizophrenia, anxiety disorders, post-traumatic stress disorder, substance use disorders, autism, and attention-deficit/hyperactivity disorder [1, 2, 3, 4, 5, 6, 7, 8, 9]. The transdiagnostic character of anhedonia suggests a common mechanism across disorders. The most systematic attempts to formalize this common mechanism have utilized concepts from reinforcement learning [10]. Early models posited that anhedonia corresponds to a reduction in reward sensitivity [11, 12], but the predictions of these models have not been consistently validated, suggesting a more complex picture [13]. Here, we argue that one neglected source of complexity is the interplay between reward sensitivity and cognitive capacity limits.

In reinforcement learning theory, states (e.g., stimuli, context) are mapped to actions by a learned policy. The amount of memory needed to store a policy is dictated by the mutual information between states and actions; any physical system (such as the brain) has a limited memory capacity. One implication of limited capacity is reward insensitivity and, thus, some aspects of anhedonia may arise from cognitive resource limitations.

Under capacity limits, policies must be *compressed* by discarding some state information [14, 15, 16]. This results in the tendency to reuse frequently chosen actions across multiple states—a form of *perseveration*, the tendency to repeat actions independently of their reinforcement history. The theory of policy compression is normative: it specifies an optimal level of perseveration for a given capacity limit. Empirically, compression strategies may differ, with some policies yielding more reward than others for the same cost. We refer to deviations from optimal perseveration as *inefficiency* because it results in a suboptimal use of finite memory (less reward for the same memory utilization). This phenotype is conceptually distinct from capacity, and can be measured separately. We argue here that capacity and efficiency may be key phenotypes for understanding cognitive disturbances in anhedonia. We show that these can be estimated from behavioral data on a widely used behavioral assay, the Probabilistic Reward Task (PRT), and that they reveal new aspects of anhedonia that would otherwise have been invisible.

We also address the underlying neural mechanisms and treatment implications. Our previous work suggested that tonic dopamine should determine the allocation of cognitive resources for task performance based on reward rate [17, 18]. Reduction in tonic dopamine should therefore produce insensitivity of task performance to reward rate [19]. It stands to reason that increasing tonic dopamine should increase reward sensitivity. We demonstrate that this is consistent with the effects of *κ* opioid receptor (KOR) antagonism, which elevates tonic dopamine [20, 21, 22, 23, 24]. We find that efficiency also increases, suggesting that tonic dopamine may not only determine the amount of resources available but also the efficiency of their allocation. Mechanistically, this might be implemented through dopamine-dependent changes in learning rate for perseveration. Finally, we find that anhedonia is associated with changes in efficiency but not memory, highlighting the clinical utility of distinguishing these computational phenotypes.

## Results

### Policy complexity and efficiency in anhedonia after *κ*-opioid receptor antagonism

We performed a secondary analysis of an 8-week, multicenter, placebo-controlled, double-blind, randomized trial to test the effects of KOR antagonism for anhedonia (Figure 1A) [25, 26]. Because this trial identified a significant treatment effect of KOR antagonism for anhedonia (as measured by the Snaith-Hamilton Pleasure Scale, SHAPS), we sought to understand the cognitive basis of this improvement. We analyzed a total of 55 participants (KOR antagonist group: *N* = 24; placebo group: *N* = 31) who completed both a baseline and post-treatment Probabilistic Reward Task (PRT). Owing to previously-reported baseline differences in anhedonia between the two groups (mean SHAPS ± SD: placebo 33.03 ± 5.54; KOR 37.29 ± 8.89, *p* = 0.0338), we analyzed the pre-treatment groups separately.

**Figure 1:**
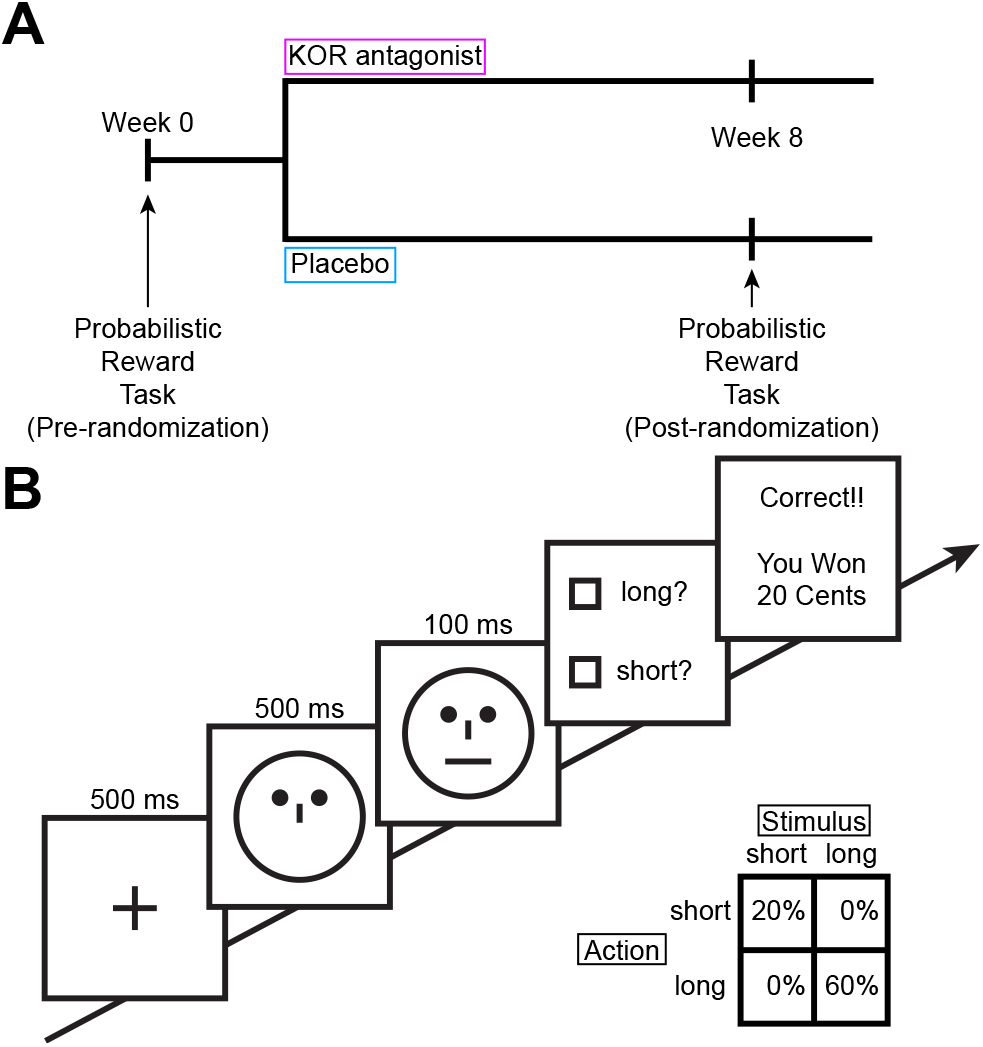
Trial and task design. A) Participants were randomized to 8 weeks of placebo (N = 31) or a KOR antagonist (N = 24) and completed the PRT at baseline and at week 8. B) On each trial of the PRT, participants fixated on a cross, followed by the presentation of a face without a mouth, followed by either a short (11.5mm) or long (13mm) mouth in the face. Participants responded by pressing one of two keyboard keys and completed 200 trials in two blocks of 100 trials. The bottom right shows an example reward schedule where the long stimulus is rewarded more often than the short stimulus. The mapping between response, stimulus, and reward was counterbalanced between participants.

The PRT is a reward-based decision making task that asks participants to discriminate two similar stimuli (Figure 1B) [27, 28]. Unbeknownst to participants, one of the two stimuli yields reward more often than the other when correctly identified. According to the theory of policy compression [16], performance in this task (average reward) depends on the amount of information participants encode about the underlying state (i.e., the stimulus identity), quantified by the mutual information between states and actions—a participant’s *policy complexity*. Each participant is assumed to have a capacity limit (upper bound on policy complexity), which delimits their achievable performance. If participants maximally utilize their capacity, their average reward should fall along an optimal reward-complexity frontier, as shown in Figure 2A,B. In the PRT, maximal reward can be obtained at a policy complexity of 1 bit, corresponding to a policy that perfectly discriminates the two stimuli. At the other extreme, a subject with no capacity will generate a policy that ignores the stimuli entirely. Participant policies tend to lie close to the optimal frontier, indicating that they are utilizing most of their capacity. At the low end of the policy complexity range, participant policies fall off the optimal frontier (Figure 2F,G), indicating under-utilization of resources (inefficiency)—a pattern also observed in previous studies [15, 29].

**Figure 2:**
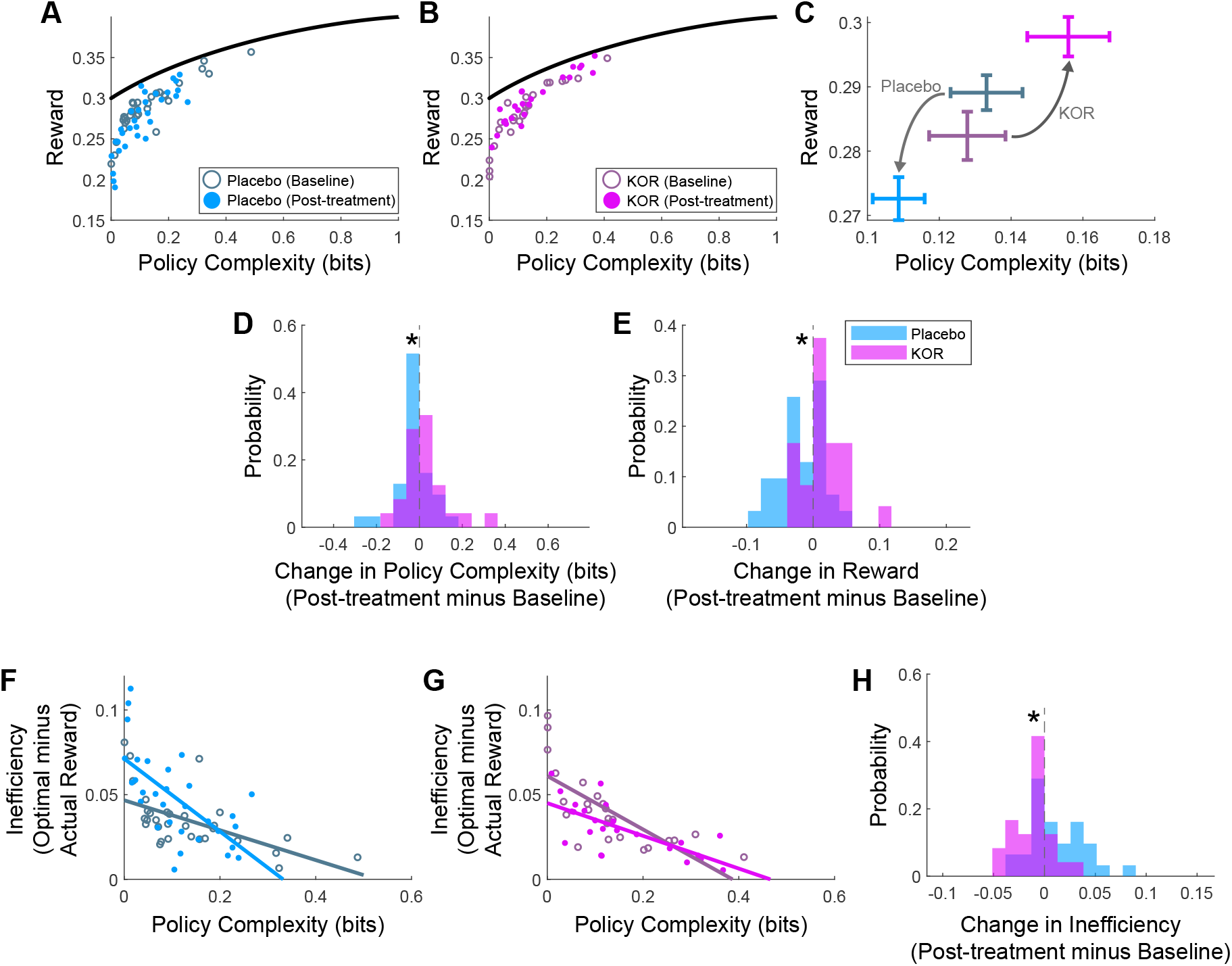
Changes in policy complexity and efficiency as a function of KOR antagonism. A,B) Reward-complexity relationship for the placebo and KOR groups at baseline and post-treatment. The black line shows the reward-complexity frontier, which indicates the optimal reward as a function of policy complexity. C) Mean ± SEM reward-complexity relationship as a function of treatment (placebo or KOR antagonism) and time (baseline or post-treatment). D) Change in policy complexity (post-treatment minus baseline) as a function of treatment. E) Change in reward (post-treatment minus baseline) as a function of treatment. F,G) Relationship between inefficiency and complexity for the placebo and KOR groups. Overlaid lines are from a linear mixed-effects model fitting inefficiency as a function of policy complexity, treatment, and time. H) Change in inefficiency (post-treatment minus baseline) as a function of treatment.

At 8 weeks, placebo treatment resulted in a decrease in both policy complexity and reward, while KOR antagonism yielded an increase in both (Figure 2C). This resulted in significant between-group differences for both policy complexity (Figure 2D; mean change in policy complexity (post-treatment minus baseline) ± SEM: placebo, -0.0245 ± 0.0141; KOR, 0.0281 ± 0.0211, *p* = 0.0362) and reward (Figure 2E; mean change in reward (post-treatment minus baseline) ± SEM: placebo, -0.0165 ± 5.61 × 10^−3^; KOR, 0.0154 ± 6.53 × 10^−3^, *p* = 4.81 × 10^−4^). Following treatment, the KOR group also became significantly more efficient compared to the placebo group (Figure 2H; mean change in inefficiency (post-treatment minus baseline) ± SEM: placebo, 0.0130 ± 4.80 × 10^−3^; KOR, -0.0109 ± 4.04 × 10^−3^, *p* = 5.68 × 10^−4^). Thus, KOR antagonism increases average reward through a combination of increasing both policy complexity and efficiency.

Policy compression makes the additional prediction that more complex policies should result in slower response times, since the brain must inspect more bits to find a coded state [16, 18, 30]. Indeed, we found that KOR antagonism, relative to placebo, slowed participants down (mean change in response times (post-treatment minus baseline) ± SEM: placebo, -59.3ms ± 23.4; KOR, 13.6ms ± 20.4, *p* = 0.0274).

To better understand how KOR treatment changed the relationship between inefficiency and policy complexity, we fit a linear mixed effects model predicting inefficiency as a function of policy complexity, treatment, and time. We identified two relevant effects: a significant treatment × time interaction (coefficient = -0.0405, *p* = 4.23 × 10^−5^), which has the effect of lowering the intercept, and a significant policy complexity × treatment × time interaction (coefficient = 0.187, *p* = 1.76 × 10^−3^), which has the effect of increasing the slope. The combination of the change in intercept and slope has the net effect of increasing efficiency as a function of policy complexity, revealing that KOR treatment increases efficiency independent of its changes to complexity. We will develop this insight further with our reinforcement learning modeling. Overall, these results suggest two orthogonal effects of KOR treatment: increases in complexity and increases in efficiency. Stated another way, participants gain increased cognitive resources *and* make better use of those resources.

### Reinforcement learning model of KOR antagonism

We developed a cost-sensitive reinforcement learning model to gain insight into how KOR antagonism affects decision making. We adapted a *Q*-learning model, ubiquitous in the reinforcement learning literature [31]. This model estimates the expected reward associated with each action for each stimulus (called *Q*-values) and updates these estimates by learning from the outcome (presence or absence of reward). Since the optimal policy under policy compression contains a marginal action probability term to engender perseveration (state-independent actions), we augmented our model with a marginal action probability term that was similarly estimated on a trial-by-trial basis. Our model contained a reward learning rate, *α*_learn_, to govern the learning of action values, a perseveration learning rate, *α*_persev_, to govern the learning of the marginal action probability, and a reward sensitivity parameter, *β*, that determines the balance between action values and perseveration in driving behavior. The *β* parameter is linked to capacity, where higher capacity is associated with higher values of *β*. Given the structure of our model, *β* is equivalent to a parameter scaling reward magnitude, as has been posited in anhedonia [12].

To model the effects of treatment, we allowed KOR and placebo to scale these parameters. Based on formal model comparison (Extended Data Table 1), we selected a model that separately scaled the perseveration learning rate, *α*_persev_, and the reward sensitivity, *β*, as a function of treatment. We confirmed that our model could recover *α*_persev_ and *β*, the parameters of interest (Extended Data Table 2). To provide confidence in the ability of our model to capture key characteristics of the data, we first fit the model to participant data and then had the model perform the PRT (using the parameter estimates for each participant) to generate a synthetic dataset (Extended Data Figure 1). This simulated dataset captured all key features of our data (see Supplementary information).

Having confirmed that our model could generate realistic data and recover parameters of interest, we turned our attention to parameter estimates to better understand how treatment affected decision making. We found that placebo and KOR antagonism scaled the perseveration learning rate, *α*_persev_ in opposite directions (Figure 3A; posterior 95% credible interval; placebo, -2.96 to -0.82; KOR, 0.61 to 1.96). The difference between KOR antagonism and placebo corresponds to the net effect of treatment on *α*_persev_, which was positive and excluded 0, showing that treatment increases perseveration (Figure 3B; difference in posterior 95% credible interval (KOR minus placebo), 1.77 to 4.32). We similarly found that placebo and KOR antagonism scaled the reward sensitivity, *β*, in opposite directions (Figure 3C; posterior 95% credible interval; placebo, -0.143 to -0.050; KOR, 0.037 to 0.138), with a treatment effect that was positive and excluded 0 (Figure 3D; difference in posterior 95% credible interval (KOR minus placebo), 0.114 to 0.254).

**Figure 3:**
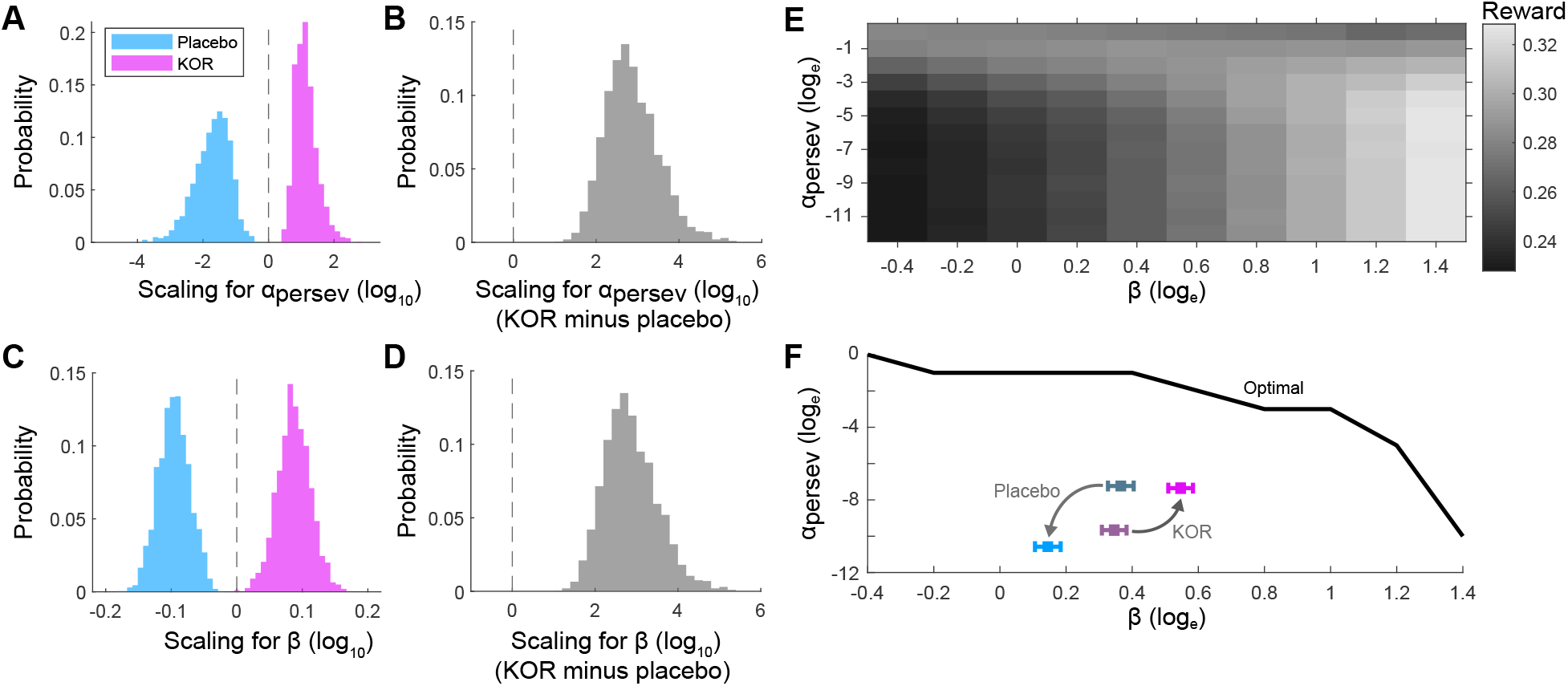
Scaling of reinforcement learning parameters as a function of KOR antagonism. A) Posterior distribution of parameter values for scaling of *α*_persev_ as a function of treatment. Scaling is multiplicative, where values greater than 0 indicate that treatment increases the parameter value, whereas values less than 0 indicate that treatment decreases the parameter value. B) Posterior distribution of treatment effect for scaling *α*_persev_, estimated as the difference in scaling between KOR and placebo. C) Posterior distribution of parameter values for scaling of *β* as a function of treatment. D) Posterior distribution of treatment effect for scaling *β*. E) Heatmap showing mean reward obtained as a function of *α*_persev_ and *β*. F) Effect of treatment in parameter space. Black line shows the optimal *α*_persev_ for each value of *β*.

To gain insight into how scaling these parameters affects decision making, we simulated datasets where we only changed parameters of interest (Extended Data Figure 1; Extended Data Table 3). Increasing only *α*_persev_ produces an increase in efficiency and a small decrease in policy complexity. The increase in efficiency manifests as a change in the intercept, but not the slope, of the relationship between inefficiency and policy complexity. Increasing only *β* produces a relatively large increase in policy complexity, which is consistent with the theoretical link between larger *β* and increased capacity. It also produces an increase in efficiency for low-complexity policies. Increasing both *α*_persev_ and *β*, like we find for KOR antagonism, produces both an increase in policy complexity and an increase in efficiency. The increase in efficiency manifests as a change in both the intercept (decrease) and the slope (increase) of the relationship between inefficiency and policy complexity, like our empirical findings.

We gained insight into the relationship between KOR antagonism and optimal behavior by visualizing the relationship between *α*_persev_, *β*, and reward, while holding *α*_learn_ fixed (Figure 3E). As *β* increases, for the optimal *α*_persev_, the net reward obtainable also increases, consistent with our theory linking higher *β* to higher capacity and higher capacity to greater reward. We also find that increasing perseverative learning is most beneficial at lower values of *β* (i.e., lower capacity), consistent with the idea that perseveration is increasingly optimal as subjects become more resource limited. In Figure 3F, we can see that the effect of KOR antagonism is to shift both *α*_persev_ and *β* closer to an optimal regime. A notable finding is the increased *α*_persev_ at baseline for the placebo group relative to the KOR group. This is consistent with the baseline difference in SHAPS between these groups, with the placebo group having lower SHAPS: the larger *α*_persev_ estimates for this group is closer to the optimal regime and is consistent with less severe anhedonia.

### Policy complexity and efficiency as a function of hedonic tone

Because the original study identified a significant improvement in the SHAPS following KOR antagonism [25], we sought to identify which mechanism—increased policy complexity, increased efficiency, or both—is associated with anhedonia. We first examined the relationship between hedonic tone and reward learning in a non-clinical population. We recruited 100 participants from Amazon Mechanical Turk and implemented a version of the PRT suitable for online delivery [32]. Participants completed the SHAPS and reported a wide range of scores (mean SHAPS ± SD: 11.45 ± 6.54, range 0 to 36). We show the reward-complexity relationship in Figure 4A. For visualization purposes only, we perform a median split of participants on the basis of SHAPS.

**Figure 4:**
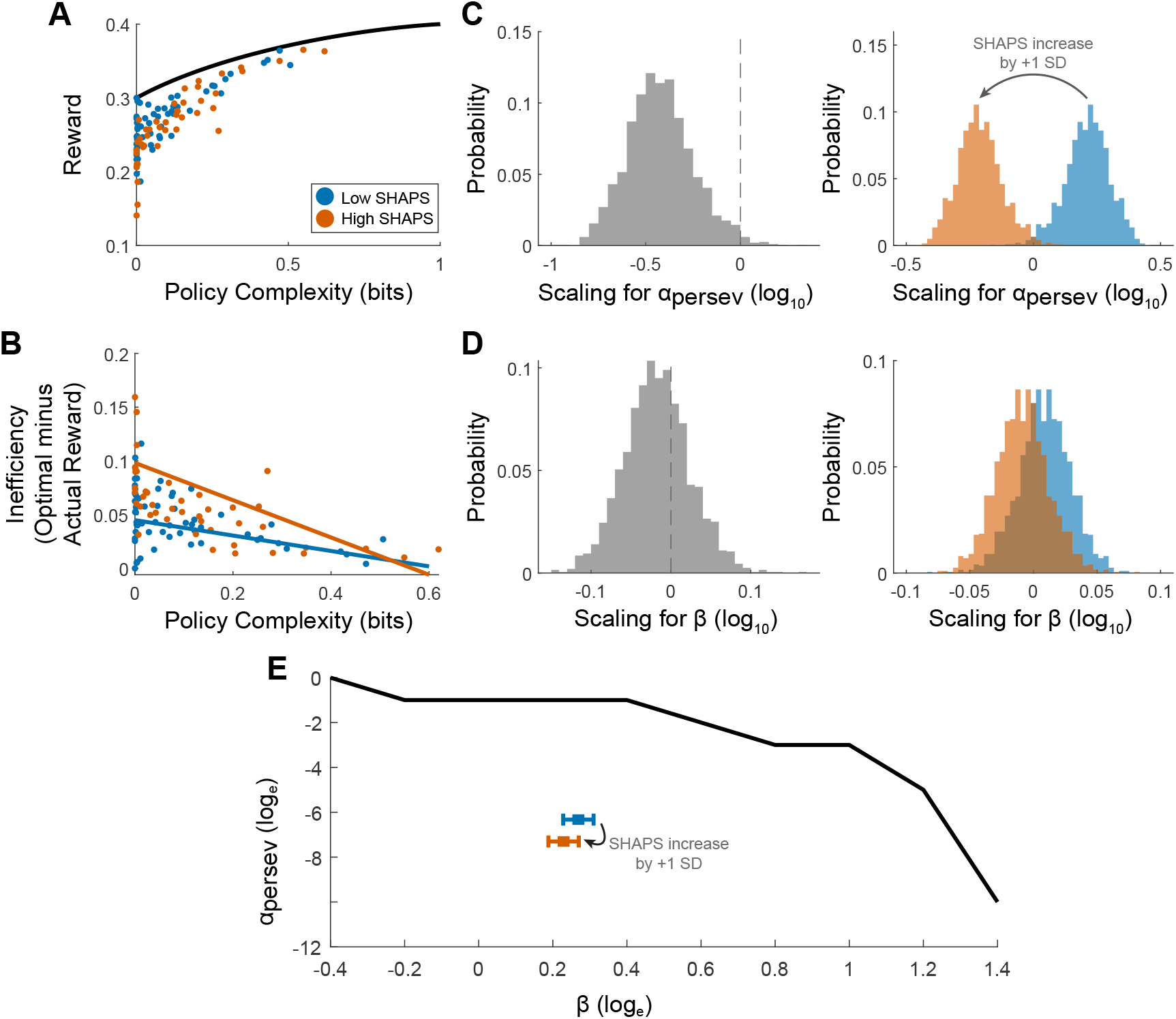
Changes in complexity, efficiency, and reinforcement learning parameters as a function of hedonic tone. A) Reward-complexity tradeoff as a function of hedonic tone. For illustration only, participants are median split on the basis of SHAPS scores into ‘Low SHAPS’ (low anhedonia) and ‘High SHAPS’ (high anhedonia). B) Inefficiency-complexity relationship as a function of hedonic tone. For illustration only, the color lines are regression fits denoting extremes of SHAPS in our dataset (blue is lowest SHAPS = 0, orange is highest SHAPS = 36). C) Left: Posterior distribution of parameter values for scaling of *α*_persev_ as a function of SHAPS. Right: An example demonstrating scaling for an increase in SHAPS of 1 SD (from 0.5 SD below the mean (blue) to 0.5 SD above the mean (orange)). D) Left: Posterior distribution of parameter values for scaling of *β* as a function of SHAPS. Right: Scaling for the same increase in SHAPS. E) Effect of variation in SHAPS in parameter space. Black line shows the optimal *α*_persev_ for each value of *β*.

Unlike the effects of KOR antagonism, we found that SHAPS did not predict policy complexity (coefficient = −5.24 × 10^−3^, *p* = 0.241). We did, however, identify a significant relationship with inefficiency. We fit a linear regression predicting inefficiency as a function of SHAPS and policy complexity and identified a significant intercept change (coefficient for effect of SHAPS = 9.55 × 10^−3^, *p* = 6.54 × 10^−3^) but not a significant slope change (coefficient for SHAPS × policy complexity interaction = −0.0182, *p* = 0.394). Given our simulations exploring the effects of changing parameters (Extended Data Figure 1), a change of intercept without a change of slope is consistent with hedonic tone affecting perseveration (*α*_persev_) and not capacity (*β*).

We reanalyzed two prior PRT datasets and found similar effects on the relationship between inefficiency and policy complexity (Extended Data Figure 2). The first was a transdiagnostic sample of patients (control group: *N* = 25; clinical group: *N* = 41, 18 with bipolar disorder, 23 with major depressive disorder) [33, 34]. These groups differed significantly in baseline anhedonia (mean anhedonic Beck Depression Inventory-II subscore ± SD: control, 0.72 ± 1.02; clinical, 5.22 ± 3.78, *p* = 2.22 × 10^−7^; mean Mood and Anxiety Symptom Questionnaire-Anhedonic Depression subscale ± SD: control, 51.5 ± 12.6; clinical, 77.1 ± 19.3, *p* = 1.45 × 10^−7^). Consistent with differences in anhedonia, when we analyzed inefficiency as a function of policy complexity and group, we identified a significant intercept difference (coefficient for clinical group = 5.29 × 10^−3^, *p* = 0.022) without a concurrent slope difference (coefficient for policy complexity × clinical group interaction = −1.71 × 10^−3^, *p* = 0.443). We additionally found no difference in policy complexity between the two groups (mean policy complexity ± SEM: control, 0.371 ± 0.043; clinical, 0.333 ± 0.024, *p* = 0.412).

The second dataset we analyzed was a test of a longstanding hypothesis relating reduced dopamine to anhedonia [35, 36]. In this double-blinded study, participants received either placebo or low-dose pramipexole—thought to reduce phasic dopamine release—and performed the PRT (placebo group: 13; pramipexole group: 11) [37]. When we analyzed inefficiency as a function of policy complexity and treatment, we identified a significant intercept effect (coefficient for treatment = 7.82 × 10^−3^, *p* = 0.043) without a significant slope effect (coefficient for policy complexity × treatment = −1.90 × 10^−3^, *p* = 0.615). We also found no difference in policy complexity as a function of treatment (mean policy complexity: placebo, 0.297 ± 0.043; pramipexole, 0.319 ± 0.057, *p* = 0.757).

### Reinforcement learning model of hedonic tone

We next fit a reinforcement learning model similar to the one we used for the KOR dataset, except now we allowed *α*_persev_ and *β* to scale as a function of SHAPS. We found that increases in SHAPS were associated with less perseveration (Figure 4C; posterior 95% credible interval: -0.739 to - 0.046). In contrast, anhedonia had no effect on modulating *β*, in contrast to KOR antagonism (Figure 4D; posterior 95% credible interval: -0.097 to 0.066). In parameter space, the net effect of an increase in SHAPS is to move participants away from an optimal regime (Figure 4E). Taken together, these data support the notion that hedonic tone spans the axis of efficiency, not capacity.

## Discussion

We leveraged a theory of resource-limited reinforcement learning to shed light on the cognitive structure of anhedonia. Building on prior work demonstrating impairments in reward sensitivity, we decomposed these impairments into separate effects of policy complexity (state-dependence of an action policy) and efficiency (utilization of cognitive resources). We found that KOR antagonism affected both of these measures, whereas anhedonia is associated only with reduced efficiency.

The finding that anhedonia is not associated with reduced complexity is surprising, in part, because complexity determines reward sensitivity, and reward insensitivity appears to be the cardinal feature of anhedonia (though see [13] for more nuance). There are a number of explanations for this apparent disconnect. One is that anhedonia may be more psychologically related to the concept of ‘liking,’ the pleasure associated with reward, rather than ‘wanting,’ the motivation furnished by reward learning [38], both of which are relevant for anhedonia. In our paradigm, reward sensitivity is related to ‘wanting,’ which would render the PRT an inappropriate assay to measure deficits in ‘liking.’ Further, the SHAPS is not designed to disambiguate these different aspects of reward processing, but newer scales such as the Dimensional Anhedonia Rating Scale [39], the Temporal Experience of Pleasure Scale [40], and the Positive Valence Systems Scale [41] provide insight into the multidimensional nature of anhedonia.

It is also plausible that anhedonia might be a consequence of reduced reward *learning*, not reduced reward sensitivity [42]. A limitation of our study is that our model could not recover the reward learning rate (Extended Data Figure 1). It is worth noting that our findings seem at odds with an influential reinforcement learning account of anhedonia implicating decreased reward sensitivity as the key causal variable [12]. Interestingly, the parameterization of that model links increased reward sensitivity with increased perseveration. Our model orthogonalizes reward sensitivity from perseveration, suggesting what was previously identified as blunted reward sensitivity may have been impaired perseverative learning (see Supplementary materials).

Under our computational framework, perseveration is closely related to habits, since habits can be similarly thought of as state-independent actions within a particular context [43]. A prediction of our findings is that anhedonia may not only manifest as a deficit in perseveration, but may also manifest as a deficit in habit formation. Intriguingly, recent work on the origin of habits has revealed that they are largely divorced from reward [44]. If true, this would highlight a cognitive deficit in anhedonia unrelated to reward processing. Altogether, our findings motivate a future research program studying habit formation in anhedonia, both important for better understanding this symptom and because it may form the basis of clinically relevant behavioral interventions.

The aspect of KOR antagonism which appears to be unrelated to anhedonia (increased policy complexity) suggests relevant clinical utility outside of anhedonia. As one example, we hypothesize KOR antagonism may prove beneficial in treating cognitive deficits in chronic schizophrenia, a clinically-relevant domain with pressing needs for psychopharmacological treatment. Cognitive deficits in schizophrenia are well-established [45] and cognitive deficits are among the strongest predictors of functional outcomes [46]. Despite decades of effort, there are no first-line pharmacotherapies for cognitive symptoms in schizophrenia [45] (though recently-developed muscarinic acetylcholine receptor agonists show promise [47, 48, 49]). We recently demonstrated that patients with chronic schizophrenia have *reduced* policy complexity relative to healthy control participants [29]. It stands to reason that increasing complexity in chronic schizophrenia, perhaps via KOR antagonism, might treat a subset of cognitive deficits and improve functional outcomes. Although it may seem counterproductive to administer dopaminergic drugs in schizophrenia, numerous studies have shown that dopamine-releasing agents can be safe to administer in this population [50, 51, 52]. Neurobiologically, our finding that KOR antagonism increases complexity is similar to our previous results following administration of dopaminergic medications in Parkinson’s disease [18]. A new subtlety of our findings here is that tonic dopamine may control the efficiency of resource allocation, a finding that is perhaps related to the role of dopamine in habit formation [53, 54, 55].

Further, anhedonia may be related to more subtle disruptions in the dopaminergic system than had been previously thought, as more global disruptions would likely reduce complexity as well.

## Conclusion

We leveraged computational principles to identify two mechanisms of action of KOR antagonism— one related to anhedonia (increase in efficiency), and one unrelated to anhedonia (increase in policy complexity). We hypothesize that the increase in complexity can be leveraged for other indications, including possibly cognitive deficits in psychosis. Our results provide a clear example of the potential for computational psychiatry to provide transdiagnostic insights that integrate across levels of analysis.

## Methods

### KOR antagonism: randomized control trial design and participants

We conducted a secondary analysis of a phase 2a clinical trial designed to test the efficacy of a novel *κ*-opioid receptor (KOR) antagonist for the treatment of anhedonia [25, 26, 56]. The trial was an 8-week, multicenter, placebo-controlled, double-blind, randomized study in a transdiagnostic sample of participants with anhedonia. Active drug was JNJ-67953964 (Aticaprant, previously CERC-501 and LY2456302), a selective KOR antagonist dosed at 10mg daily. Since this trial used a biomarker-based proof-of-mechanism approach, the preregistered primary outcome was a change in functional magnetic resonance imaging of the ventral striatum during reward anticipation, measured at baseline and 8 weeks. Preregistered secondary outcomes were a change in the mean Snaith-Hamilton Pleasure Scale (SHAPS), a clinically-validated measure of anhedonia [57], assessed every 2 weeks, and a change in the response bias - a measure of reward learning - on the Probabilistic Reward Task. The trial was preregistered at NCT02218736. We report here a secondary analysis of the Probabilistic Reward Task, which was not part of the preregistered protocol.

Participants were aged 21 to 65, recruited from six US centers, had a SHAPS of at least 20 (assessed using dimensional scoring guidelines [58]), and had a DSM-IV TR diagnosis of major depressive disorder, bipolar I or II depression, generalized anxiety disorder, social phobia, panic disorder, or post-traumatic stress disorder. Participants were enrolled after providing informed consent to a protocol approved by each local institutional review board. Our dataset for secondary analysis consisted of 55 patients (KOR antagonist group: *N* = 24 [44%]; mean age ± SD, 39.2 ± 13.9 years; 10 males [42%]; placebo group: *N* = 31 [56%]; mean age ± SD, 40.8 ± 13.7 years; 12 males [39%]) [26]).

### Non-clinical population: study design and participants

We conducted an online-based study to assess how variation in hedonic tone affects reward learning in a non-clinical population. We recruited 100 participants (mean age ± SD, 41.9 ± 11.5; 62 males [62%]) from Amazon Mechanical Turk. We selected our sample size based on an effect size we assumed would be half of what we identified for the KOR dataset (*f* ^2^ = 0.1297) with a desired power of 90% to maximize the probability of identifying an effect. These participants completed the Probabilistic Reward Task followed by a demographic survey and the SHAPS. Participants gave informed consent, and the Harvard University Committee on the Use of Human Subjects approved the experiment.

### Clinical population: study design and participants

We reanalyzed data from patient populations performing the PRT [33, 34]. The dataset consisted of 66 total participants (control group: *N* = 25 [38%]; mean age ± SD, 38.4 ± 10.8; 14 males [56%]; clinical group: *N* = 41 [62%]; mean age ± SD, 41.9 ± 10.3; 24 males [59%]; 18 with bipolar disorder [44%], 23 with major depressive disorder [56%]). The control participants had no psychiatric diagnosis and were taking no psychoactive medications. In addition to the PRT, participants completed the Beck Depression Inventory-II and the Mood and Anxiety Symptom Questionnaire.

### Pramipexole administration: study design and participants

We reanalyzed data from a double-blind, randomized trial assessing the effect of pramipexole, a D2/D3 receptor agonist, on reward learning in the PRT [37]. Participants (placebo group: 13 [54%]; mean age ± SD, 24.8 ± 3.2; 8 males [62%]; pramipexole group: 11 [46%]; mean age ± SD, 26.0 ± 5.8; 6 males [56%]) were randomized to placebo or pramipexole. In the pramipexole group, participants received a single 0.5mg dose, a low dose thought to act as a dopamine antagonist and reduce phasic dopamine release. Participants completed the PRT 2 hours after receiving placebo or pramipexole.

### Snaith-Hamilton Pleasure Scale (SHAPS)

The SHAPS is a 14-item questionnaire used to assess anhedonia across four domains: interest/pastimes, social interaction, sensory experience, and food/drink. Participants are asked to respond to pleasurable situations (e.g., I would enjoy being with my family or close friends) with one of the following responses on the basis of the last few days: strongly disagree, disagree, agree, strongly agree. According to dimensional scoring guidelines [58], scores range from 1 for strongly agree to 4 for strongly disagree, yielding a range of 14 to 56, with higher scores corresponding to greater anhedonia. The SHAPS is the only clinical measure of anhedonia that significantly changes with treatment in clinical trials [25, 59, 60].

### Probabilistic Reward Task (PRT)

The PRT is a computerized decision making task designed to elicit learning in response to reward [27, 28]. On each trial, participants observe one of two difficult-to-discriminate stimuli and are asked to report which stimulus they observed. In the clinical trial, stimuli consisted of cartoon faces with either a short mouth (11.5 mm) or a long mouth (13 mm) presented for 100 ms and participants responded by pressing one of two keyboard keys (‘z’ or ‘/’). Participants completed 200 trials in two 100 trial blocks, instead of 300 trials as usual, owing to time constraints imposed by the clinical trial [25]. In the online-based task, stimuli consisted of images of either 10 squares/7 circles or 7 squares/10 circles (with 8 variations of each) and participants reported whether they observed more squares or circles with one of two keyboard keys (‘A’ or ‘L’) [32]. Participants completed 300 trials in three 100 trial blocks. Importantly, and unbeknownst to participants, correctly responding to one stimulus yielded reward on 60% of trials (‘rich’ stimulus) while correctly responding to the other stimulus yielded reward on 20% of trials (‘poor’ stimulus). They were instructed that not all correct responses would yield a reward. The rich/poor stimuli and responses were counterbalanced across participants in both studies. For our analyses, we excluded the first 25 trials to allow behavior to stabilize. Our findings were qualitatively similar if we changed this trial exclusion threshold.

### Policy compression: a capacity limit applied to decisions

All information processing systems—the human brain included—must contend with resource limitations when making decisions. These constraints take on many forms, including computational costs [61], metabolic costs [62], interference costs [63], and others [64]. Under policy compression, we formalize the cognitive cost as the mutual information between states and actions, the policy *complexity* :

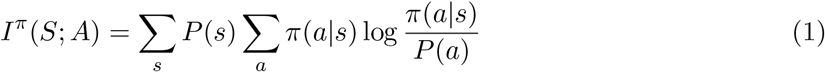

where *P* (*a*) = Σ_*s*_ *P* (*s*)*π*(*a*|*s*) is the marginal probability of choosing action *a* under the policy. In general, we assume that policies are subject to a capacity constraint, an upper bound, *C*, on policy complexity. Shannon’s noisy channel theorem states that the minimum expected number of bits to transmit a signal across a noisy information channel without error is equal to the mutual information. Therefore, if the optimal policy requires more memory than the subject possesses, then it must *compress* the policy, or render it less state-dependent. We define the optimal policy, *π*^*∗*^, as:

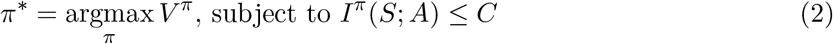

where *V* ^*π*^ is the expected reward under policy *π*:

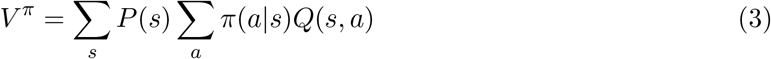

and *Q*(*s, a*) is the expected reward for taking action *a* in state *s*.

We can express our constrained optimization problem in the following unconstrained Lagrangian form:

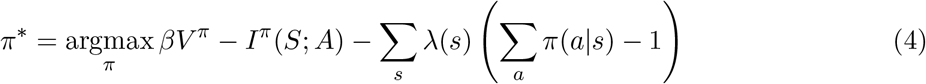

where *β* ≥ 0, *λ*(*s*) ≥ 0 are Lagrangian multipliers. Solving this equation reveals that the optimal policy takes on the following form:

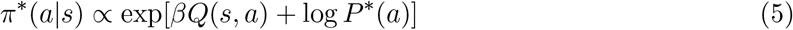

where *P*^*∗*^(*a*) is the optimal marginal action distribution, which can be interpreted as a form of perseveration.

The optimal policy takes the form of the familiar softmax distribution, common in the reinforcement learning literature. Here, the Lagrange multiplier, *β*, plays the role of the inverse temperature parameter. Note that although *β* typically takes on the role of balancing exploration/exploitation in reinforcement learning, we made no such appeals in deriving this policy. Moreover, *β* is a function of the policy complexity:

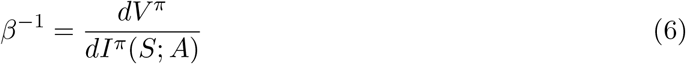

At high policy complexity, when 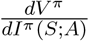 is shallow, the optimal *β* is large and the policy is dominated by *Q*-values. At low policy complexity, the optimal *β* is close to 0, and *Q*-values have minimal impact on the policy. Moreover, when *β* is small, the perseveration term, log *P*^*∗*^(*a*), dominates, and the policy is largely state-independent.

To construct the empirical reward–complexity curves, in both datasets, we computed the average reward according to equation 3, where *P* (*s*) = [0.5, 0.5] and 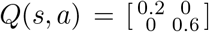, by construction, and *π*(*a*|*s*) was calculated from empirical action frequencies. We estimated mutual information by computing the empirical action frequencies for each state for each session.

### Reinforcement learning modeling

We constructed a cost-sensitive *Q*-learning model which contains three parameters (*α*_learn_, *α*_persev_, and *β*) and estimates action values, *Q*(*s, a*), and marginal action probability, *P* (*a*), to generate actions according to the following policy, mimicking the optimal policy under policy compression:

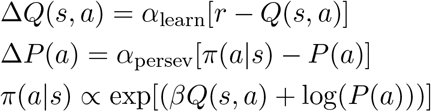

where *r* = 1 if the current trial is rewarded and 0 otherwise. The key feature of our model is a mechanism that allows treatment to multiplicatively scale *α*_persev_ and *β* (obtained after model comparison, see below). The model scales parameters in the following manner:

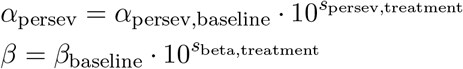

A scaling value of 0 results in no scaling, *>* 0 results in an increase, and *<* 0 results in a decrease. For our online study, we scaled parameters as a function of the *z*-scored SHAPS in the following manner:

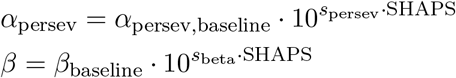

We initialized *Q*(*s, a*) at 0 and *P* (*a*) at 0.5 and we assumed scaling terms equaled 0 on sessions without treatment. We included all trials for analysis. Learning rates were constrained not to exceed 1. We constructed hierarchical models to obtain estimates of each parameter. Parameters were drawn from the following distributions:

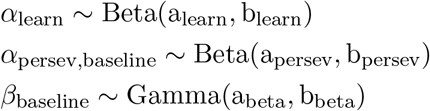

where we used Cauchy^+^(0,5) as a weakly-informative prior for each parameter. The gamma distribution was parameterized with a shape (a_beta_) and scale (b_beta_) parameter. Finally, the scaling terms were drawn according to

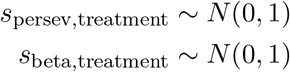

*α*_learn_, *α*_persev,baseline_, and *β*_baseline_ were constrained at the group level (one parameter per participant) and scaling terms were constrained at the treatment level (one parameter per treatment).

We initially fit a model that scaled all parameters (*s*_learn,treatment_, *s*_persev,treatment_, *s*_beta,treatment_), which produced an estimate of *s*_learn,treatment_ that did not differ from 0, suggesting that treatment did not effect *α*_learn_. We therefore compared this ‘full’ model to the ‘reduced’ model we present above (which does not scale *α*_learn_). We performed model comparison using Pareto-smoothed importance sampling leave-one-out cross-validation to estimate the expected log predictive density, a validated measure of Bayesian model evaluation [65]. We found that our reduced model produced a similar fit. We next compared our reduced model to three simpler variants: one that only scaled *α*_persev_, one that only scaled *β*, and one with no scaling of any parameters. Model comparison favored the model we present above which scales *α*_persev_ and *β* (Extended Data Table 1).

We next performed posterior predictive checks. We used the mean of each parameter as a point estimate and simulated 200 trials of the PRT for each participant to mimic the dataset that was used to fit the model. We analyzed this simulated dataset in the exact manner we analyzed the ground-truth dataset.

To provide confidence in our interpretation of parameter changes, we tested the ability of our model to recover known parameters. Using the same fictive, simulated dataset as above, we fit our reinforcement learning model and obtained recovered parameter estimates. We computed Pearson’s correlation between the known and recovered parameters (Extended Data Table 2).

For our heatmap of reward obtained with different *α*_persev_ and *β* combinations, we ran 3,000 independent simulations of the PRT for each combination of parameters. We fixed *α*_learn_ at 0.1423, the mean posterior estimate across all participants. We performed a grid search across thirteen logarithmically-spaced *α*_persev_ values from *e*^−12^ to *e*^0^, and ten *β* values from *e*^−0.4^ to *e*^1.4^.

Models were fit using R 4.2.2 (accessed with RStudio 2022.12.0+353) using the Rstan package (version 2.26.13). We performed model comparison using the loo package (version 2.5.1).

## Data Availability

The KOR dataset is available in the National Institute of Mental Health (NIMH) Data Archive repository and can be accessed by contacting the NIMH Data Archive at the following email address. Other data included in the present study are available upon reasonable request to the authors.

## Statistical analyses

For all group-level differences, we computed two-sided *t*-tests. In the KOR dataset, owing to repeated measures, we fit linear mixed effects models to predict 1) inefficiency and 2) the probability of choosing the richer option. Independent variables were policy complexity, treatment (placebo or KOR), and time (baseline or post treatment), with a random intercept per participant. For the other datasets (online non-clinical, clinical, and pramipexole), we fit a linear regression to predict inefficiency. For the online non-clinical dataset, the dependent variables were policy complexity and *z*-scored SHAPS. For the clinical dataset, they were policy complexity and group (control or clinical). For the pramipexole dataset, they were policy complexity and treatment (placebo or pramipexole). All analyses were 2-sided with an *α* of 0.05.

## Extended data

**Extended Data Figure 1:**
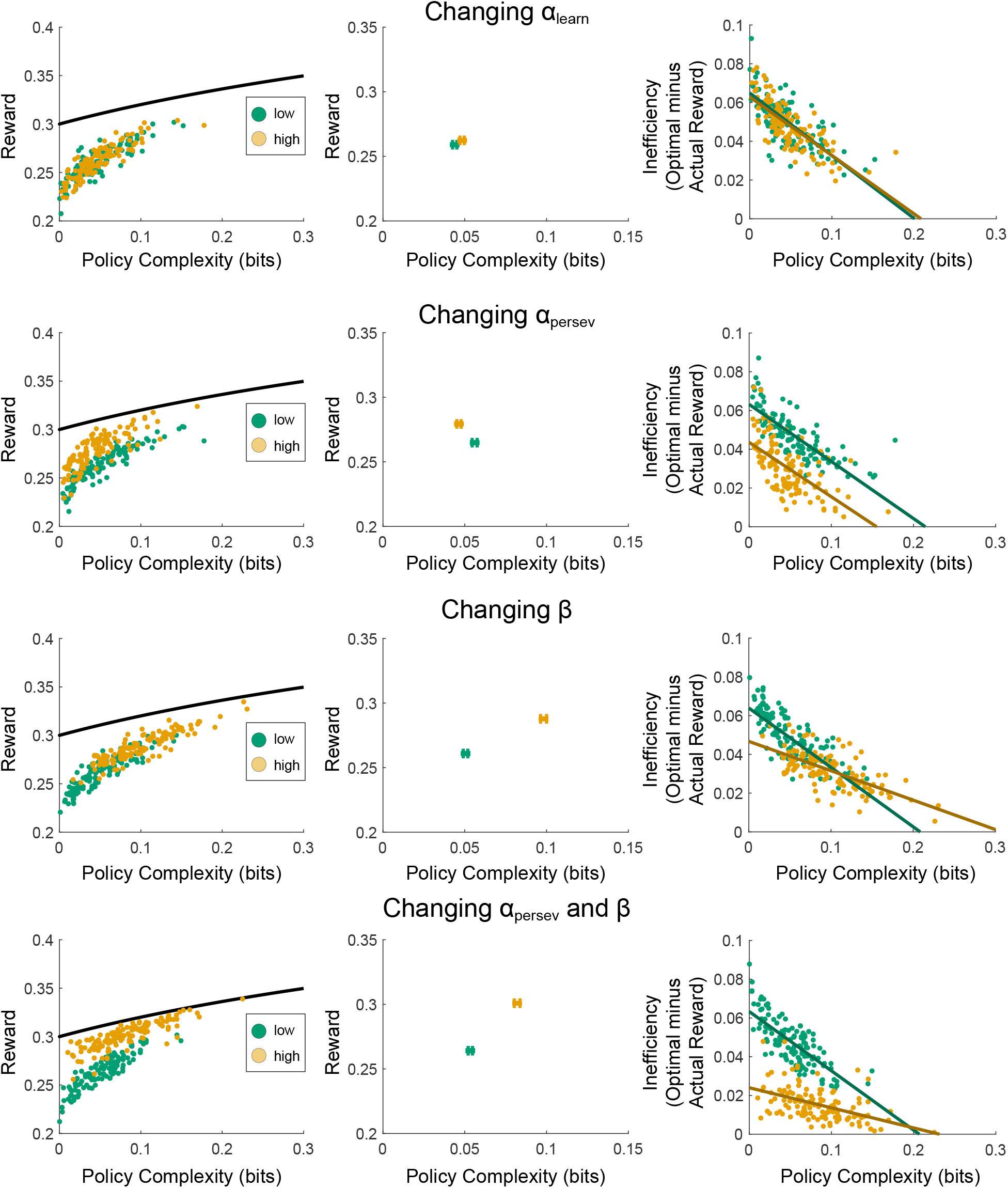
Effect of changing reinforcement learning model parameters on reward-complexity relationship and inefficiency. Parameter values for simulation are given in Extended Data Table 3.

**Extended Data Figure 2:**
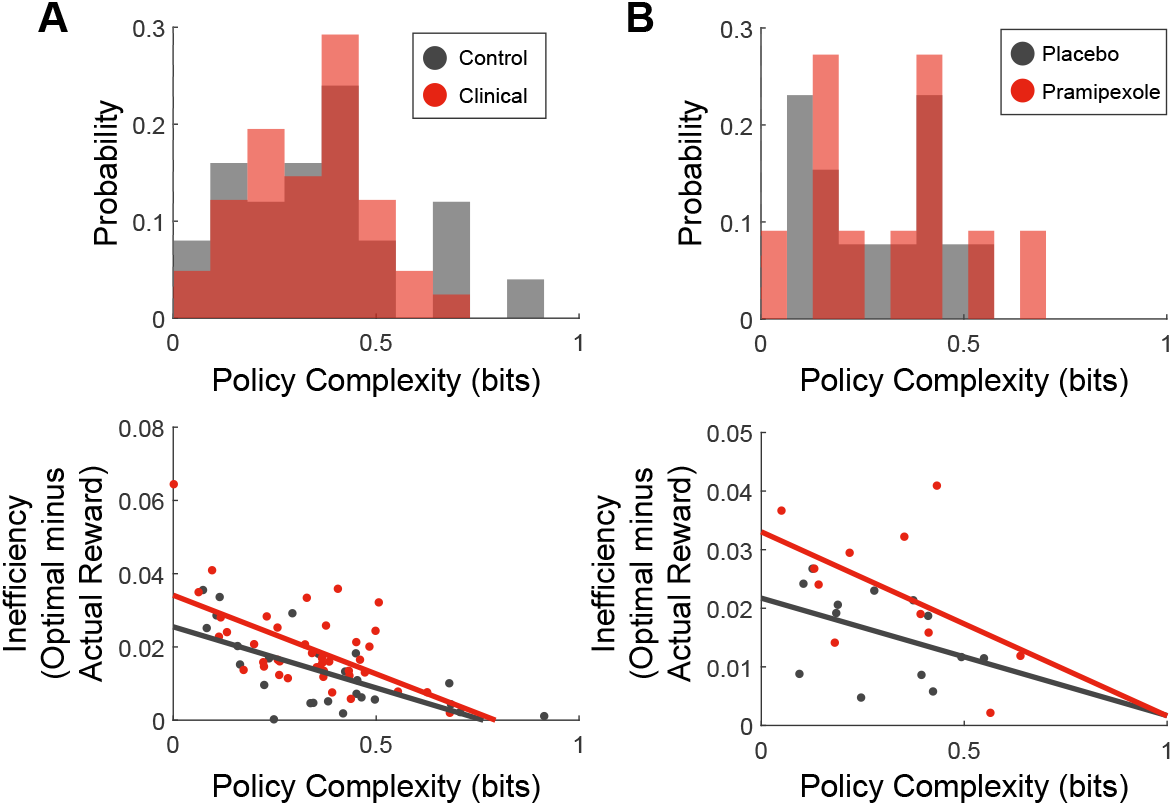
Policy complexity and inefficiency for reanalyzed PRT datasets. A) Clinical dataset from [33] and [34]. B) Pramipexole dataset from [37].

**Extended Data Table 1:** Model comparison using Pareto-smoothed importance sampling leave-one out cross validation. A difference in the expected log predictive density of 4 points provides evidence in favor of a model. The first model, which scales *α*_persev_ and *β*, is favored over the second, which scales all parameters, since it provides similar expected predictive accuracy with fewer parameters.

| Model | | | Expected log predictive<br>density difference ± SE | Effective number<br>of parameters (p_loo) ± SE |
| --- | --- | --- | --- | --- |
| Scale $\alpha_{\text{learn}}$ | Scale $\alpha_{\text{persev}}$ | Scale $\beta$ | | |
| | x | x | $0.0 \pm 0.0$ | $100.5 \pm 1.2$ |
| x | x | x | $-1.8 \pm 2.5$ | $107.6 \pm 1.3$ |
| | x | | $-14.7 \pm 6.1$ | $112.9 \pm 1.4$ |
| | | x | $-29.2 \pm 7.8$ | $100.8 \pm 1.1$ |
| | | | $-43.1 \pm 10.1$ | $98.5 \pm 1.1$ |

**Extended Data Table 2:** Parameter recovery.

| Parameter | Pearson Correlation Between<br>Actual and Recovered Parameter<br>(95% CI) |
| --- | --- |
| $\alpha_{\text{learn}}$ | 0.412 (0.183 - 0.607) |
| $\alpha_{\text{persev}}$ | 0.941 (0.862 - 0.983) |
| $\beta$ | 0.926 (0.884 - 0.952) |

**Extended Data Table 3:** Parameters used for Extended Data Figure 1 simulations.

| Simulation | Low Parameter | High Parameter | Fixed Parameters |
| --- | --- | --- | --- |
| Changing $\alpha_{\text{learn}}$ | $\alpha_{\text{learn}} = 0.09$ | $\alpha_{\text{learn}} = 0.18$ | $\alpha_{\text{persev}} = 2.5 \cdot 10^{-4}$<br>$\beta = 1.4$ |
| Changing $\alpha_{\text{persev}}$ | $\alpha_{\text{persev}} = 2.5 \cdot 10^{-4}$ | $\alpha_{\text{persev}} = 2.5 \cdot 10^{-2}$ | $\alpha_{\text{learn}} = 0.14$<br>$\beta = 1.4$ |
| Changing $\beta$ | $\beta = 1.4$ | $\beta = 2.2$ | $\alpha_{\text{learn}} = 0.14$<br>$\alpha_{\text{persev}} = 2.5 \cdot 10^{-4}$ |
| Changing $\alpha_{\text{persev}}$ & $\beta$ | $\alpha_{\text{persev}} = 2.5 \cdot 10^{-4}$<br>$\beta = 1.4$ | $\alpha_{\text{persev}} = 2.5 \cdot 10^{-2}$<br>$\beta = 2.2$ | $\alpha_{\text{learn}} = 0.14$ |

## Supplementary information

### Reinforcement learning model of KOR antagonism: behavioral simulations on the Probabilistic Reward Task

Treatment increased policy complexity (Supplementary Figure 1D; mean change in policy complexity (post-treatment minus baseline) ± SEM: placebo, -0.0214 ± 7.69 × 10^−3^; KOR, 0.0269 ± 8.32 × 10^−3^, *p* = 9.07 × 10^−5^) and reward (Supplementary Figure 1E; mean change in reward (post-treatment minus baseline) ± SEM: placebo, -0.0201 ± 4.36 × 10^−3^; KOR, 0.0199 ± 4.62 × 10^−3^, *p* = 7.21 × 10^−8^). There was a significant decrease in inefficiency (Supplementary Figure 1H; mean change in inefficiency (post-treatment minus baseline) ± SEM: placebo, 0.0161 ± 3.50×10^−3^; KOR,-0.0149 ± 3.53 × 10^−3^, *p* = 1.08 × 10^−7^). Using the same linear mixed effects model to predict inefficiency as a function of policy complexity, treatment, and time, we found a significant treatment × time interaction (coefficient = -0.0346, *p* = 1.94 × 10^−7^) and a significant policy complexity × treatment × time interaction (coefficient = 0.290, *p* = 8.66 × 10^−4^).

**Supplementary Figure 1:**
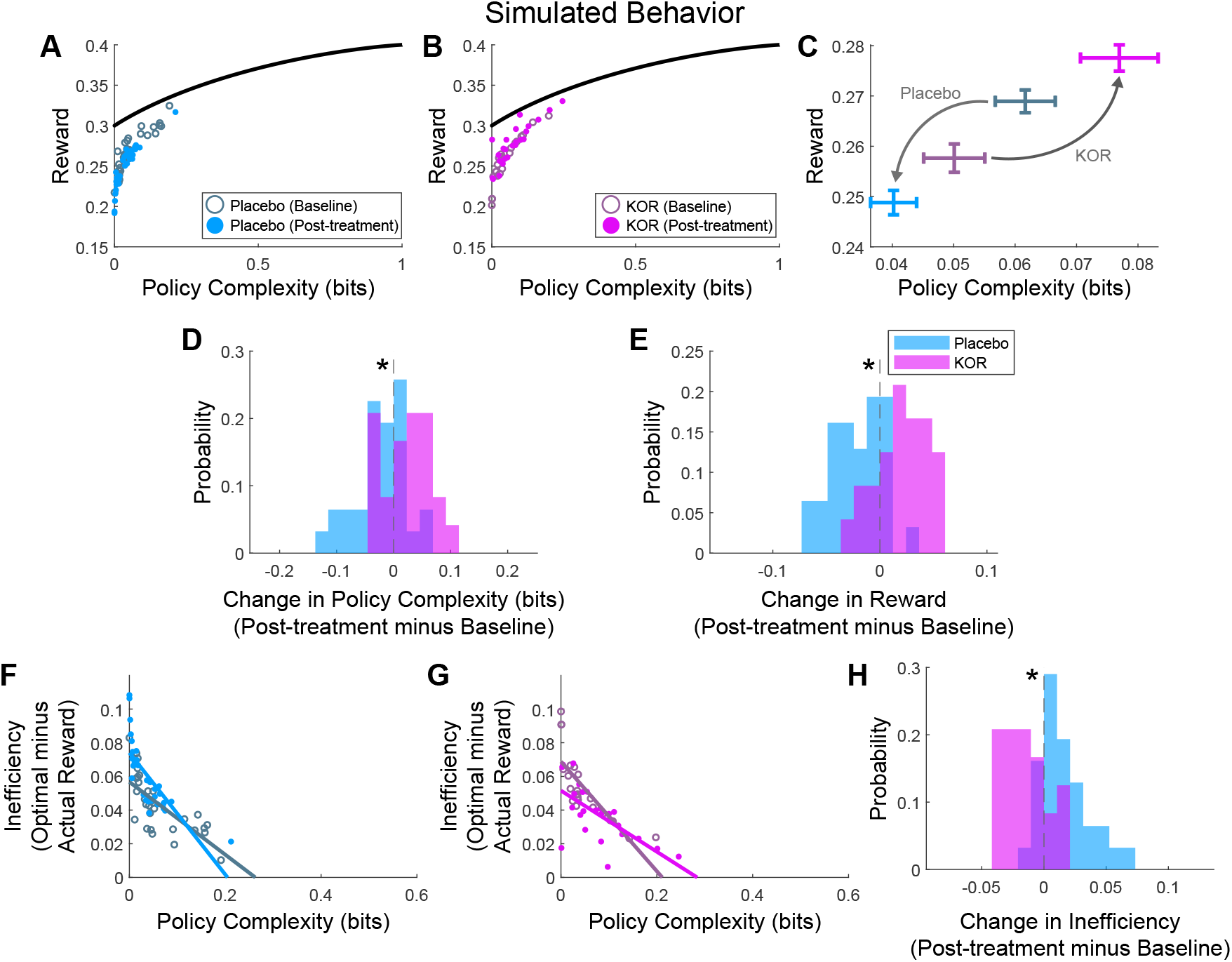
Simulation: changes in complexity and efficiency as a function of KOR antagonism. A,B) Reward-complexity relationship for placebo and KOR groups, at baseline and post-treatment. C) Mean ± SEM reward-complexity relationship as a function of treatment and time. D) Change in policy complexity as a function of treatment. E) Change in reward as a function of treatment. F-G) Inefficiency-complexity relationship for placebo and KOR groups. H) Change in inefficiency as a function of treatment.

### Anhedonia model from Huys et al (2013)

Huys et al (2013) developed a reinforcement learning model, fit to PRT data, describing anhedonia as a reduction in reward sensitivity [12]. We will show that the parameterization of reward sensitivity in this model produces a similar effect as our perseveration term.

In their model, reward prediction errors are computed by scaling binary reward, *r*, by a reward sensitivity parameter *ρ*. These reward prediction errors are multiplied by *ϵ*, the learning rate, to iteratively update *Q*-values.

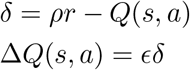

Given the reward structure in the PRT, this has the effect of scaling *Q*-values by *ρ* as 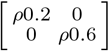.

These *Q*-values are used to update choice weights, which are fed through a standard softmax decision rule to generate a policy:

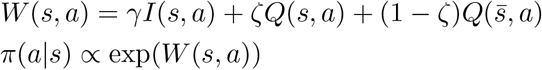

The choice weights of this model contain two noteworthy components. The first i s a n instruction variable, *I*(*s, a*), where *I*(*s, a*) = 1 for the instructed action for a given stimulus, and 0 otherwise. Instructions are scaled by *γ* to capture how strongly instructions influence c hoice. T he second component describes sensory ambiguity and allows *Q*-values for the non-presented stimulus – 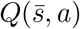- to ‘leak’ into the policy. This is done by the *ζ* parameter, where *ζ* ∈ [0.5, 1]; *ζ* = 1 describes no sensory ambiguity (only *Q*(*s, a*) contributes) and *ζ* = 0.5 describes complete sensory ambiguity (*Q*(*s, a*) and 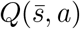 contribute equally).

To see how this sensory ambiguity rule leads to perseveration, we can define *ζ* = *θ* + 0 .5, where *θ* ∈ [0, 0.5] and replace *ζ* in the choice weights:

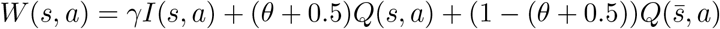

which we can rearrange as

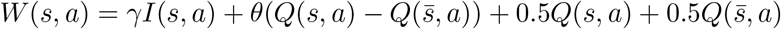

Since the states are equiprobable (*p*(*s*) = 0.5), this latter set of terms,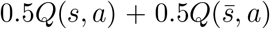, can be written as 𝔼_*s*_(*Q*(*s, a*)) = Σ_*s*_ *p*(*s*)*Q*(*s, a*), the expected *Q*-value of taking action *a*. We can therefore write the weights as

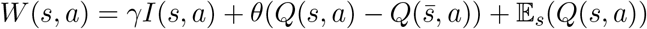

Written this way, weights are a function of three variables: 1) *I*(*s, a*), the instructions, 2) 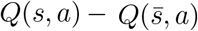, the difference in *Q*-values between the observed and non-observed states, to account for sensory ambiguity, and 3) 𝔼_*s*_(*Q*(*s, a*)), a state-independent value term which can be thought of as a kind of perseveration since it will generate an action bias.

We ran a simulation to gain an intuition into how 𝔼_*s*_(*Q*(*s, a*)) engenders perseveration (Supplementary Figure 2). In this simulation, 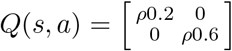, meaning 𝔼_*s*_(*Q*(*s, a*)) ∝ *ρ*0.6 for the richer option and ∝ *ρ*0.2 for the leaner option. Intuitively, 𝔼_*s*_(*Q*(*s, a*)) will proportionally favor the richer option as reward sensitivity grows, leading to an action bias.

**Supplementary Figure 2:**
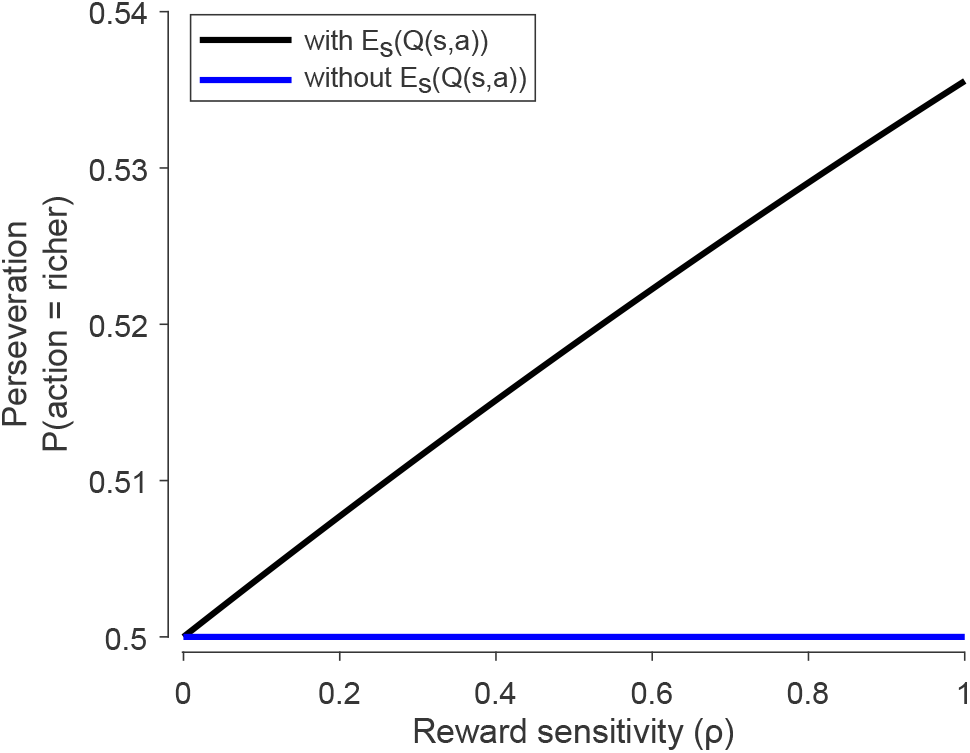
Increasing reward sensitivity (*ρ*) in the Huys et al (2013) model leads to perseveration. Increasing reward sensitivity (*ρ*) leads to increased perseveration (black). To demonstrate that the 𝔼_*s*_(*Q*(*s, a*)) term is responsible for perseveration, we ran the same simulation with the 𝔼_*s*_(*Q*(*s, a*)) removed (blue). For this simulation, we used *γ* = 1 and *ϵ* = 0.25. Findings were insensitive to choice of *γ* and *ϵ*.

